# Validation of a two-stage automated screening pipeline for medical systematic reviews: a stratified agreement study with three independent human reviewers, using PABAK and Gwet’s AC1

**DOI:** 10.64898/2026.10.05.26364147

**Authors:** Enrico Pirani, Davide Costazza, Pier Paolo Berti

## Abstract

**Background:** Large language models (LLMs) are increasingly used to automate title–abstract screening in systematic reviews, but validation studies typically report sensitivity, specificity, or Cohen’s kappa — metrics that are unstable under the low and unbalanced prevalence that characterises most reviews. No previous study has simultaneously combined a medical topic; a hybrid two-stage pipeline (rules + LLM); a three-way classification with a separately sampled uncertain class; stratified sampling with three independent human reviewers; and dual reporting of prevalence-robust coefficients (PABAK and Gwet’s AC1).

**Objective:** To validate a two-stage automated screening pipeline (a rule-based Python script followed by a reproducible LLM adjudication of uncertain records) against three independent human reviewers in a medical systematic review on high on-treatment platelet reactivity (HTPR), aspirin resistance, and carotid plaque. The primary validation target is the Stage 1 automatic-exclusion decision (670/904 records), which is the workload the pipeline is designed to take off the human reviewer.

**Methods:** From 904 screened records (Stage 1: 131 INCLUDE, 670 EXCLUDE, 94 UNCERTAIN; Stage 2 LLM via API, temperature = 0: 11 INCLUDE, 83 EXCLUDE), a stratified sample of 185 records (50 per Stage 1 stratum + 35 Stage 2; seed = 42) was judged independently and blind to the automated labels by three physicians: E.P. (pipeline designer), D.C. and P.P.B. (both external to the thesis project). Concordance was quantified with sensitivity, specificity, PABAK (*q* = 3 throughout), Gwet’s AC1, work saved over sampling (WSS), and number needed to read (NNR) — AI versus each reviewer and between reviewers — overall and by stratum. Majority labels (2 of 3) are reported as a supplementary panel; they do not replace pairwise tables as the primary reference for sensitivity.

**Results:** On the primary target, Stage1-EXCLUDE concordance was PABAK = AC1 = 1.000 versus E.P. (48/48) and versus D.C. (50/50), and PABAK = 0.940 / AC1 = 0.959 versus Berti (48/50 EXCLUDE, 2 UNCERTAIN). That stratum is 670/904 records (74.1%). Pairwise human agreement was highest between the two external readers (Costazza vs Berti: PABAK = 0.854, AC1 = 0.881) and lower when E.P. was one of the pair (0.703 / 0.774 vs Costazza; 0.621 / 0.702 vs Berti). AI versus each human remained poor (PABAK 0.225 / 0.205 / 0.222). Majority labels on 185 records were 146 EXCLUDE / 27 UNCER-TAIN / 4 INCLUDE / 8 NO_MAJORITY. The machine labelled INCLUDE 0 of the 4 majority-INCLUDE records (74, 75, 76, 80). Overall sensitivity was 0.800 versus E.P. (8/10; false negatives 623, 712), 0.500 versus D.C. (1/2; false negative 76), and 0.833 versus Berti (5/6; false negative 76). Versus majority the false-negative count is 1 (record 76); records 623 and 712 are majority EXCLUDE. Stage1-INCLUDE sensitivity of 1.000 is tautological. WSS = 74.1% describes the exclusion stratum only.

**Conclusions:** What this study validates, now with three independent physicians, is automatic exclusion at Stage 1. Fine classification of INCLUDE records is not validated: humans do not agree on which residual records are INCLUDE (triple INCLUDE intersection = record 74 only), Stage 2 missed most of the few human INCLUDE labels it received, and the machine recovered none of the four majority-INCLUDE records. Domain-familiarity as an explanation of INCLUDE divergence remains a hypothesis, not a measured variable.

## 1 Introduction

Systematic reviews (SRs) are a cornerstone of evidence-based medicine, yet their production is increasingly constrained by the exponential growth of the biomedical literature. Title–abstract screening accounts for a substantial share of the total workload and is typically performed by at least two independent human reviewers (Borah et al., 2017; Page et al., 2021). The manual nature of this step creates both a time bottleneck and a source of human error, with disagreements documented even among experienced assessors (Wang et al., 2020).

Large language models (LLMs) have emerged as a plausible route to partial automation of this process, operating in zero-shot or few-shot mode without task-specific training (Scherbakov et al., 2025; Galli et al., 2025). The performance of automated screening tools is, however, highly sensitive to the choice of validation metric.

The most widely used measure of screening concordance, Cohen’s kappa, is mathematically unstable under the prevalence conditions typical of SRs (Byrt et al., 1993; Feinstein and Cicchetti, 1990). Prevalence-robust alternatives — PABAK and Gwet’s AC1 — remain uncommon, although they have been reported (Guo et al., 2024; Zink et al., 2024; Nawrath et al., 2026; Rodríguez et al., 2026; Gwet, 2008). A second under-discussed issue is the handling of borderline records; a third is the validity of the comparison when the designer of the pipeline is also a human validator.

We present a prospective validation of a two-stage automated pipeline applied to a medical systematic review on HTPR, aspirin resistance, and carotid plaque, with a design that explicitly addresses two-stage architecture, three-way classification, stratified sampling, and three independent human reviewers. We report PABAK, Gwet’s AC1, and — in response to the Research Synthesis Methods editorial guidance for evaluating ML/GenAI tools in SRMA (Farotimi et al., 2026) — the standard workload-reduction metrics (work saved over sampling, number needed to read).

## 2 Related work

A growing number of studies have evaluated LLMs for title–abstract screening. Among the 101 studies retrieved by a systematic search (Section 3.1), four reported both PABAK and Gwet’s AC1: Guo et al. (2024), Zink et al. (2024), Nawrath et al. (2026), and Rodríguez et al. (2026). None of those four combined a medical two-stage rules-plus-LLM pipeline, a separately sampled uncertain class, stratified dual or triple human review, and WSS/NNR in the same design. Tao et al. (2025, LitAutoScreener) combined a twostage pipeline with stratified sampling, but with binary classification and a non-standard metric. No study among the 101 scored 5/5 on the coded dimensions in File S3.

## 3 Methods

### 3.1 Search strategy for related work

Four complementary strategies: findpapers v0.6.7 (PubMed + Scopus); federated MCP search across 14 sources; citation chaining via Semantic Scholar; manual addition. After deduplication: 508 unique records, reduced to 101 after heuristic screening. Full details are in File S1.

### 3.2 Study design and context

Prospective agreement study validating a two-stage automated pipeline applied to a medical systematic review on HTPR, aspirin resistance, and carotid plaque. Dataset: 904 records from PubMed, Scopus, Europe PMC, OpenAlex, and arXiv.

### 3.3 The two-stage screening pipeline

#### Stage 1 — rule-based script

A Python script (no machine-learning or language-model component) applied keyword heuristics derived from the PICO model, assigning INCLUDE, EXCLUDE, or UNCER-TAIN. The rules are specific to the HTPR / aspirin-resistance / carotid-plaque domain. Result: 131 INCLUDE (14.5%), 670 EXCLUDE (74.1%), 94 UNCERTAIN (10.4%).

#### Stage 2 — LLM adjudication

The 94 UNCERTAIN records were submitted to DeepSeek V4 Pro (model identifier: `deepseek-chat`) via a reproducible API call (temperature = 0, JSON `response_format`, fixed prompt). API access was in August 2026; no response JSON was retained, so the calendar day of the canonical call is not recoverable. Three successive API writes were made on the same 94 records (0/94, then 3/91, then 11/83 INCLUDE/EXCLUDE). The third write is the result reported here and in File S4. A prior non-API conversational pass (27/67) is not the reported result. The prompt underwent a pilot calibration phase before the final version reported here: the Population and Outcome criteria were checked against the official Methods chapter of the review protocol (explicitly including ischaemic stroke as an alternative population to documented carotid plaque, and the pharmacogenetic association as a valid outcome); a judgement rule calibrated to the level of evidence available in an abstract was introduced after a pilot test on a subset of records had revealed an anomalous exclusion rate attributable to full-text-level stringency being incorrectly applied to abstract-level screening. In the final version, anchor criteria (population, core of the intervention) require plausible evidence; detail criteria (comparator design, specific numeric thresholds) may be deferred to full text. Final result: 11 INCLUDE (11.7%) and 83 EXCLUDE (88.3%) of the 94 UNCERTAIN records. The complete prompt, including the history of versions tested, is reported in File S5. Labels in the Stage 2 slice of File S4 match this API file on all 35 sampled records (3 INCLUDE: 35, 215, 900; 32 EXCLUDE).

### 3.4 Sampling design for validation

Stratified sample of 185 records: 50 from each of the three Stage 1 strata (seed = 42) + 35 from Stage 2. The stratified design was preferred to simple random sampling given the dominant size of the EXCLUDE stratum (~74.1%). The sample is a subset of the full dataset (904 records); results for the Stage1-EXCLUDE stratum were used to estimate (not directly measure) work saved over sampling on the full population (Section 3.6).

### 3.5 Human reference standard: three independent reviewers

Validation was conducted by three physicians, each blind to the automated decisions and to the other two readers: Enrico Pirani (E.P., author of the Stage 1 script and of the Stage 2 prompt), Davide Costazza (D.C., external to the thesis project and not involved in pipeline design), and Pier Paolo Berti (P.P.B., external to the thesis project, same written PICO sheet, scored in blind). E.P. completed 182/185 judgements (missing 213, 708, 890); D.C. and P.P.B. completed all 185. P.P.B. is a third independent rater of the full sample, not an adjudicator of pairwise disagreements. Disagreements were not resolved into a forced consensus list.

The reviewers were selected with an explicit asymmetry by design. E.P. has prior familiarity with the corpus as author of the thesis project from which this systematic review originates. D.C. and P.P.B. are external to that project. Blindness to per-record AI labels does not make E.P. independent of the classification logic he authored; only the two external readers are independent of that logic. Inter-reviewer concordance is therefore reported separately for the primary decision (exclude / do not exclude, Sections 4.4–4.6) and the secondary decision (which residual records are INCLUDE, Section 5.1).

Majority labels (agreement of at least two of the three readers) are computed for a supplementary panel. Eight records have three different labels (NO_MAJORITY) and are kept as their own category; they are not recoded as UNCERTAIN. Majority is not the primary reference for sensitivity.

Risk of training/test contamination. DeepSeek V4 Pro was used in zero-shot mode, with no training or fine-tuning on the present study’s data: there is therefore no data-leakage risk in the conventional supervised-learning sense. It cannot, however, be excluded that some records in the dataset (published before the model’s training cut-off) were present in the LLM’s pretraining corpus; this is an intrinsic and not fully mitigable limitation of any zero-shot validation that uses published literature, and it is declared here in accordance with transparency guidance for manuscripts evaluating GenAI tools in SRMA (Farotimi et al., 2026).

### 3.6 Concordance and practical-impact metrics

Sensitivity and specificity: INCLUDE as the positive class; UNCERTAIN records excluded from the binary analysis.

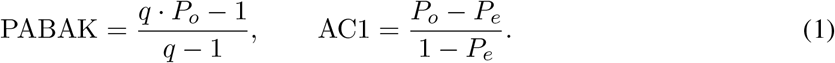

Thresholds: *<* 0.40 poor; 0.40–0.75 moderate; *>* 0.75 excellent. 95% confidence intervals by the Wilson method. All PABAK cells use *q* = 3.

In response to the Research Synthesis Methods editorial guidance for ML/GenAI evaluation manuscripts in SRMA (Farotimi et al., 2026), we additionally report two standard practical-impact metrics: Work Saved over Sampling (WSS), estimated as the proportion of records excluded by Stage 1 with validated sampled specificity, extrapolated from the sample estimate to the full population of 904 records; and Number Needed to Read (NNR) = 1*/*precision, computed for the Stage1-INCLUDE stratum.

### 3.7 Stratified analysis

All metrics were computed separately for each of the four sampling strata and for each pairwise comparison (AI vs each human; humans vs each other). Custom Python script (`agreement_metrics.py`, File S2), fully open-source and available in the data repository (Data availability section).

## 4. Results

### 4.1 Literature gap analysis

None of the 101 identified studies simultaneously satisfied the five methodological dimensions of the present design (Table 1). Four studies reported PABAK and AC1 together; none of those four also combined the remaining four dimensions.

**Table 1:** Closest related studies, scored on the five design dimensions of the present study. Scores in File S3. No study among the 101 reached all five dimensions. The present row is the design under test, not an external comparator.

| Study | Domain | Two-stage | 3-way | Strat. | PABAK+AC1 | Dual/triple |
| --- | --- | --- | --- | --- | --- | --- |
| Tao et al., 2025 | Medicine | Yes | No | Yes | No | No |
| Guo et al., 2024 | Medicine | No | No | No | Yes | No |
| Zink et al., 2024 | Medicine | No | No | No | Yes | No |
| Nawrath et al., 2026 | Environment | No | No | No | Yes | No |
| Rodríguez et al., 2026 | Rehabilitation | No | No | No | Yes | No |
| Present study | Medicine | Yes | Yes | Yes | Yes | Yes (3) |

**Table 2:** Decision distributions for the pipeline and for each reviewer. Majority INCLUDE ids: 74, 75, 76, 80. NO_MAJORITY ids: 1, 9, 20, 61, 142, 213, 261, 612. Record 213 is incomplete (E.P. missing), not a three-way clash of complete votes.

| Source | $n$ judged | INCLUDE | EXCLUDE | UNCERTAIN | Other |
| --- | --- | --- | --- | --- | --- |
| Pipeline AI (full sample) | 185 | 53 (28.6%) | 82 (44.3%) | 50 (27.0%) | — |
| E.P. (reviewer 1) | 182 | 13 (7.1%) | 165 (90.7%) | 4 (2.2%) | 3 missing |
| D.C. (reviewer 2) | 185 | 5 (2.7%) | 151 (81.6%) | 29 (15.7%) | — |
| P.P.B. (reviewer 3) | 185 | 9 (4.9%) | 137 (74.1%) | 39 (21.1%) | — |
| Majority (2 of 3) | 185 | 4 (2.2%) | 146 (78.9%) | 27 (14.6%) | 8 NO_MAJORITY (4.3%) |

**Table 3:** Pairwise concordance (*q* = 3).

| Comparison | $n$ | $P_o$ | PABAK | AC1 | Reading |
| --- | --- | --- | --- | --- | --- |
| AI vs E.P. | 182 | 48.4% | 0.225 | 0.315 | Poor |
| AI vs D.C. | 185 | 47.0% | 0.205 | 0.278 | Poor |
| AI vs Berti | 185 | 48.1% | 0.222 | 0.277 | Poor |
| E.P. vs D.C. | 182 | 80.2% | 0.703 | 0.774 | High on EXCLUDE; not on INCLUDE |
| E.P. vs Berti | 182 | 74.7% | 0.621 | 0.702 | Moderate |
| D.C. vs Berti | 185 | 90.3% | 0.854 | 0.881 | Excellent (two external readers) |

**Table 4:**
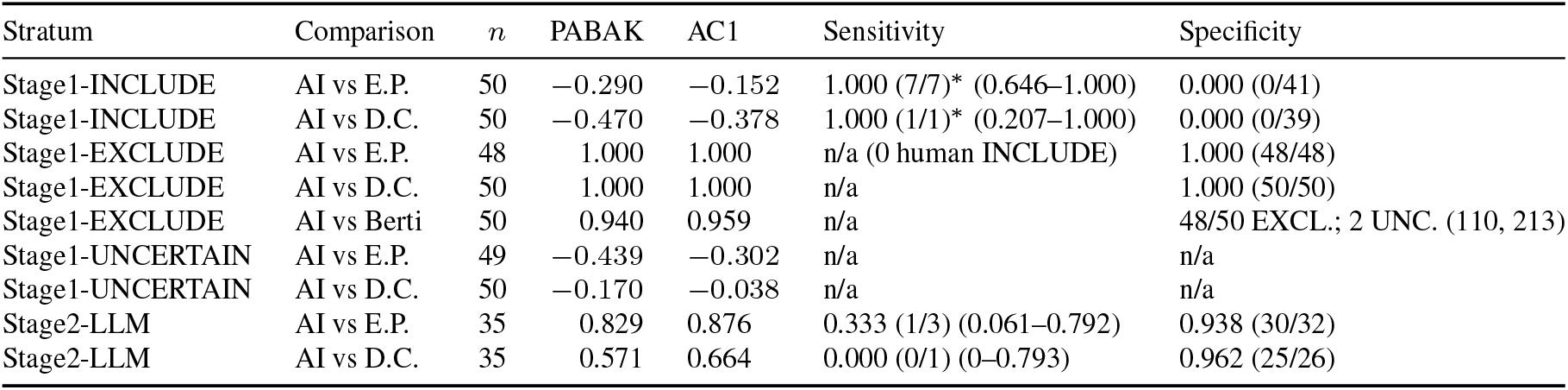
Concordance by sampling stratum. PABAK uses *q* = 3 in every cell. Stage1-INCLUDE sensitivity of 1.000 is tautological. Wilson 95% CIs in parentheses where a binary INCLUDE count exists.

| Stratum | Comparison | $n$ | PABAK | AC1 | Sensitivity | Specificity |
| --- | --- | --- | --- | --- | --- | --- |
| Stage1-INCLUDE | AI vs E.P. | 50 | -0.290 | -0.152 | 1.000 (7/7)* (0.646–1.000) | 0.000 (0/41) |
| Stage1-INCLUDE | AI vs D.C. | 50 | -0.470 | -0.378 | 1.000 (1/1)* (0.207–1.000) | 0.000 (0/39) |
| Stage1-EXCLUDE | AI vs E.P. | 48 | 1.000 | 1.000 | n/a (0 human INCLUDE) | 1.000 (48/48) |
| Stage1-EXCLUDE | AI vs D.C. | 50 | 1.000 | 1.000 | n/a | 1.000 (50/50) |
| Stage1-EXCLUDE | AI vs Berti | 50 | 0.940 | 0.959 | n/a | 48/50 EXCL.; 2 UNC. (110, 213) |
| Stage1-UNCERTAIN | AI vs E.P. | 49 | -0.439 | -0.302 | n/a | n/a |
| Stage1-UNCERTAIN | AI vs D.C. | 50 | -0.170 | -0.038 | n/a | n/a |
| Stage2-LLM | AI vs E.P. | 35 | 0.829 | 0.876 | 0.333 (1/3) (0.061–0.792) | 0.938 (30/32) |
| Stage2-LLM | AI vs D.C. | 35 | 0.571 | 0.664 | 0.000 (0/1) (0–0.793) | 0.962 (25/26) |

### 4.2 Pipeline output

Stage 1: 131 INCLUDE (14.5%), 670 EXCLUDE (74.1%), 94 UNCERTAIN (10.4%) of 904 records.

Stage 2 (reproducible API call): 11 INCLUDE (11.7%) and 83 EXCLUDE (88.3%) of the 94 UNCER-TAIN records.

### 4.3 Characteristics of the validation sample

Stratified sample of 185 records. E.P. completed 182/185 judgements (missing records 213, 708, and 890); D.C. and P.P.B. completed all 185. Observed INCLUDE counts: E.P. 13, D.C. 5, Berti 9, pipeline 53.

### 4.4 Overall pairwise concordance

All PABAK values use *q* = 3. UNCERTAIN labels from either rater were dropped from sensitivity and specificity, as pre-specified in Section 3.6.

AI vs E.P. (*n* = 182): *P*_*o*_ = 48.4%; PABAK = 0.225 (poor); AC1 = 0.315 (poor); sensitivity = 0.800 (8/10); specificity = 0.645 (78/121). False negatives versus E.P.: records 623 and 712 (Stage 2; pipeline EXCLUDE, E.P. INCLUDE).

AI vs D.C. (*n* = 185): *P*_*o*_ = 47.0%; PABAK = 0.205 (poor); AC1 = 0.278 (poor); sensitivity = 0.500 (1/2); specificity = 0.652 (75/115). False negative versus D.C.: record 76.

AI vs Berti (*n* = 185): *P*_*o*_ = 48.1%; PABAK = 0.222 (poor); AC1 = 0.277 (poor); sensitivity = 0.833 (5/6); specificity = 0.723 (73/101). False negative versus Berti: record 76.

Inter-human concordance:

- E.P. vs D.C. (*n* = 182): *P*_*o*_ = 80.2%; PABAK = 0.703; AC1 = 0.774. Joint EXCLUDE 141/182. INCLUDE intersection 1 (record 74); union 17.
- E.P. vs Berti (*n* = 182): *P*_*o*_ = 74.7%; PABAK = 0.621; AC1 = 0.702. INCLUDE intersection 1 (record 74).
- D.C. vs Berti (*n* = 185): *P*_*o*_ = 90.3%; PABAK = 0.854; AC1 = 0.881. INCLUDE intersection 4 (74, 75, 76, 80); union 10.

These facts are not interchangeable. Concordance on the exclude/do-not-exclude decision — the primary validation target — is high, and in the Stage1-EXCLUDE stratum is complete or near-complete across three readers. Concordance on which of the remaining records are INCLUDE is not. The IN-CLUDE intersection of all three humans is record 74 only. A plausible structural reading, not tested in this design, is unequal prior familiarity with the HTPR corpus. The data do not separate that familiarity difference from intrinsic ambiguity in the eligibility construct, nor from the fact that E.P. wrote the rules. Jaccard coefficients on the INCLUDE class are reported in Section 4.5 as descriptive overlap, not as evidence for that reading.

### 4.5 Concordance by stratum and INCLUDE-class overlap

On the INCLUDE class specifically, pairwise set overlap from File S4 with Berti is:

- AI vs E.P.: 8*/*58 = 0.138 (seven Stage1-INCLUDE records — 9, 20, 61, 123, 136, 261, 612 — plus Stage 2 record 900)
- AI vs D.C.: 1*/*57 = 0.018
- AI vs Berti: 5*/*57 = 0.088
- E.P. vs D.C.: 1*/*17 = 0.059
- E.P. vs Berti: 1*/*21 = 0.048
- D.C. vs Berti: 4*/*10 = 0.400

Those coefficients describe class-positive overlap. They are not a recovery rate, and they are not evidence that domain familiarity explains the INCLUDE mismatch.

### 4.6 Majority panel (supplementary) and false negatives

Majority (2 of 3) on 185 records: 146 EXCLUDE, 27 UNCERTAIN, 4 INCLUDE (74, 75, 76, 80), 8 NO_MAJORITY (4.3%). Seven of the eight NO_MAJORITY records sit in Stage1-INCLUDE.

The machine labelled those four majority-INCLUDE records UNCERTAIN, UNCERTAIN, EXCLUDE, UNCERTAIN. Intersection of pipeline INCLUDE with majority INCLUDE is empty.

False negatives (pipeline EXCLUDE, human INCLUDE):

- versus E.P.: 623, 712
- versus D.C.: 76
- versus Berti: 76
- versus majority: 76 only

Records 623 and 712 are majority EXCLUDE (both external readers agree with the machine, not with E.P.). The change from two false negatives versus E.P. to one versus majority is a change of referent, not evidence that the pipeline improved.

Costazza versus majority, after dropping NO_MAJORITY, is tautological on this sample (177/177): Costazza is never the lone dissenting vote when a majority forms, because majority is largely Costazza + Berti. That comparison is not reported as an independent replica.

### 4.7 Practical-impact metrics: WSS and NNR

#### Work saved over sampling (WSS)

Automatic exclusion of the Stage 1 EXCLUDE slice: 670 of 904 records (74.1%). In the validation sample of that stratum, E.P. 48/48, D.C. 50/50, Berti 48/50 EXCLUDE plus 2 UNCERTAIN (no INCLUDE). The ratio 670/904 describes how much of the source list the rule-based filter removed, anchored to three independent sampled readings. It is not a statement about pipeline-wide recall. The pipeline as a whole produced false negatives in Stage 2. Extrapolation from *n* = 48–50 to the remaining unsampled EXCLUDE records assumes within-stratum homogeneity, which this design cannot test.

#### Number needed to read (NNR)

In Stage1-INCLUDE, after dropping human UNCERTAIN labels, precision versus E.P. is 7*/*48 = 0.146 (NNR = 6.9) and versus D.C. is 1*/*40 = 0.025 (NNR = 40.0). Fine positive classification is the part of screening that this study does not show to be delegable.

## 5 Discussion

### 5.1 Principal findings

The primary validation target is the Stage 1 automatic-exclusion decision: 670 of 904 records (74.1%). Three physicians working blind to the labels and to each other confirmed that slice (perfect or near-perfect three-way EXCLUDE; Berti’s two UNCERTAIN labels are not INCLUDE). That result is the central empirical contribution.

Overall AI–human concordance remains poor because the validation sample over-represents INCLUDE and UNCERTAIN relative to the source list. Overall sensitivity is 0.800 versus E.P., 0.500 versus D.C., and 0.833 versus Berti, because Stage 2 missed human INCLUDE labels (record 76 on both external readers; 623 and 712 only versus E.P.). Stage1-INCLUDE sensitivity of 1.000 is tautological.

Inter-human coefficients are high on EXCLUDE and not on INCLUDE. The only record labelled INCLUDE by all three humans is 74; the machine left it UNCERTAIN. The two external readers agree with each other more than either agrees with E.P. (PABAK 0.854 versus 0.703 and 0.621). That pattern is compatible with a familiarity difference. It is also compatible with E.P. having written the rules. Familiarity was not pre-measured. The pattern is reported; it is not promoted to a confirmed mechanism (Limit 10).

### 5.2 Choice of concordance and impact metrics

PABAK and Gwet’s AC1 were chosen because Cohen’s kappa is unstable at the prevalence typical of systematic-review screening (Byrt et al., 1993; Gwet, 2008). Following Farotimi et al. (2026), WSS and NNR were added as workload metrics attached to the strata that generate them. All PABAK cells use *q* = 3.

### 5.3 The UNCERTAIN category and three reviewers as a design feature

The three reviewers used UNCERTAIN at different rates (E.P. 2.2%; D.C. 15.7%; Berti 21.1%). Dual, then triple, review was introduced to test whether results depended on a single human. For automatic exclusion they do not. For fine positive classification they do.

Stage 2 missed two of E.P.’s three Stage-2 INCLUDE labels and the external readers’ Stage-2 INCLUDE label (record 76). The uncertain class is the place where the pipeline’s false negatives appear. That is a limit on Stage 2, not on the Stage 1 exclusion filter.

### 5.4 Limitations

First, the validation is specific to one medical systematic review and to one Stage 1 script. Nothing in the design tests transport to another PICO.

Second, the Stage 1 UNCERTAIN rate (10.4%) is an output of these particular heuristics.

Third, the Stage 2 validation subsample (*n* = 35) produces sensitivity confidence intervals that are too wide to support a performance claim for the LLM stage.

Fourth, the Stage 2 model was not varied across providers or versions.

Fifth, the third reader is an independent rater of the full sample, not an adjudicator of pairwise disagreements. Eight records (4.3%) have no majority. No forced consensus INCLUDE list exists.

Sixth, WSS = 74.1% is an extrapolation from a sample of 48–50 Stage1-EXCLUDE records to 670 records.

Seventh, residual training/test contamination for Stage 2 cannot be excluded for records that may have been in the LLM’s pretraining corpus (Section 3.5).

#### Eighth — prompt calibration on the validation corpus

The Stage 2 prompt was calibrated iteratively on the same 94 UNCERTAIN records that constitute the Stage 2 population. Three successive API writes on that set produced 0/94, then 3/91, then 11/83 INCLUDE/EXCLUDE. The anchor-versus-detail rule was introduced between those writes. The reported inclusion rate is 11.7% (11/94). A stratified sample of 35 of those 94 records later entered the validation set. There is no held-out calibration set.

#### Ninth — designer as validator

E.P. designed the Stage 1 rules and the Stage 2 prompt and then served as a validator. Results that rest only on E.P. cannot be read as independent confirmation. The Stage1-EXCLUDE result does not rest only on E.P.: two external readers reproduced it.

#### Tenth — unmeasured difference in corpus familiarity

E.P. has direct prior familiarity with this literature. D.C. and Berti are external to the thesis project. The design did not record, scale, or control that difference. Two independent external readers now exist, and they agree with each other more than with E.P. That is a second data point compatible with a familiarity reading. It is not a confirmatory test of Limit 10. *n* = 2 external readers; familiarity was not pre-measured. Future studies should record each reviewer’s prior domain familiarity before scoring begins.

### 5.5 Implications

For clinicians: on this HTPR review the filter can remove the obviously off-topic papers (~3/4 of the list). Three physicians, two of them outside the project, agree on those “no”. Doubtful and candidate-yes papers still need a human. The machine is not a replacement for screening. The result does not travel automatically to another PICO.

For methodologists: pairwise tables remain the primary analysis when a majority of three is available. Majority is a useful supplementary panel and a way to count how often three readers cannot form a majority (here 4.3%). PABAK above 0.70 can coexist with an INCLUDE triple intersection of one record. Prompt development should be separated from the validation corpus. Reviewer familiarity with the source domain should be a recorded design variable.

## 6 Conclusions

This study compared a two-stage automated screening pipeline with three independent human reviewers on a stratified sample of 185 records drawn from a 904-record medical systematic review. PABAK and Gwet’s AC1 were computed with *q* = 3 throughout. WSS and NNR were computed from the strata that define them. Majority labels are supplementary.

The primary result is the Stage 1 automatic-exclusion decision. Three physicians confirmed that slice (E.P. 48/48; D.C. 50/50; Berti 48/50 EXCLUDE + 2 UNCERTAIN). The stratum accounts for 670 of 904 source records (74.1%). That is the workload the pipeline is designed to take off the reviewer, and that is what the study validates. The figure is not a claim that the pipeline as a whole has recall of 1.000.

The secondary results concern the residual task. Stage 2 sensitivity is low. Stage1-INCLUDE sensitivity of 1.000 is tautological and coexists with specificity of 0.000. The machine labelled INCLUDE 0 of 4 majority-INCLUDE records. Inter-human coefficients are high on EXCLUDE and not on INCLUDE (triple intersection = record 74). A plausible, untested reading of that mismatch is unequal prior familiarity with the corpus. E.P. designed the pipeline he then scored. The Stage 2 prompt was calibrated on the same 94-record corpus that later contributed to validation.

Automatic exclusion at Stage 1, in this PICO, is the result that three independent medical reviewers support. Fine classification of INCLUDE records remains a human, domain-experienced task, not shown here to be delegable.

## Data Availability

All data underlying the concordance analyses are provided as supplementary files with this preprint (Files S1-S5, the three-rater validation table, and agreement_metrics.py). A persistent-identifier deposit (Zenodo or OSF) will be created at journal submission. GitHub is a working copy, not the archival record.

https://github.com/Enrico68/academic-workspace/tree/main/llmscreen-validation

## Author contributions

E.P.: conceptualisation, design of the automated pipeline, data analysis, human validation (reviewer 1), drafting of the manuscript. D.C.: independent human validation (reviewer 2), critical revision of the manuscript. P.P.B.: independent human validation (reviewer 3), critical revision of the manuscript. All authors approved the final version.

## Declaration of competing interests

The authors declare no financial or non-financial competing interests relating to this work.

## Data availability

A persistent-identifier deposit (Zenodo or OSF) of Files S1–S5, File S4 with Berti majority labels, and `agreement_metrics.py` will be created at submission; the DOI will replace this placeholder. GitHub is not the archival copy. Per journal policy, data are not “available on request”.

Until the DOI is issued, the versioned files used for this manuscript are those accompanying the submission and the tagged snapshot of the public repository `Enrico68/academic-workspace` (folder `llmscreen-validation/`). Licence intended: CC-BY 4.0 for data; code under the repository licence.

## Funding

The authors received no specific funding for this work from public, commercial, or not-for-profit agencies.

## Acknowledgements

The authors declare the use of the following artificial-intelligence tools. DeepSeek V4 Pro (DeepSeek AI; model identifier `deepseek-chat`; temperature = 0; API access August 2026, calendar day not recoverable) executed Stage 2 adjudication; that use is described in Section 3.3. Claude Sonnet 4.6 (Anthropic) and Grok 4.6 (xAI) were used for language editing, structural revision, and independent recalculation of concordance statistics from the raw CSV files during manuscript preparation (September 2026). Kimi K2 (Moonshot AI) was used for full-text comparison of related work. No AI tool is an author. The scientific content, the human validation judgements, and final editorial responsibility are the authors’.

## Supplementary materials

File S1. Search strategy.

File S2. `agreement_metrics.py`.

File S3. `gap_analysis_CORRECTED.csv` (101 studies). File S4. Human validation data, two-rater table.

File S4b. Same sample with Berti decisions and majority column (185 records). File S5. Structured Stage 2 prompt, with history of versions tested.

## Notes

### Competing Interest Statement

The authors have declared no competing interest.

