## Supplementary Materials (Files S1-S5) for "Validation of a two-stage automated screening pipeline for medical systematic reviews: a stratified agreement study with three independent human reviewers, using PABAK and Gwet’s AC1": File_S1_search_strategy_EN.pdf

### Supplementary File S1. Search strategy for the literature-gap analysis

English version of the search log. The Boolean strings were already in English. Search date: 3 July 2026. Studies published after that date are not in the gap analysis.

#### 1. Objective

To identify previous studies that validated large-language-model (LLM) tools for title-abstract screening in systematic reviews, and to place the present study against that literature on five design dimensions: application domain, pipeline architecture, classification granularity, validation sampling design, and reported agreement metric.

#### 2. Retrieval (four complementary sources)

##### 2.A Structured Boolean search (findpapers v0.6.7)

Databases: PubMed and Scopus. Four Boolean queries, run on 3 July 2026.

| Query | Boolean string | Focus |
|---|---|---|
| A | (LLM OR GPT OR "generative AI") AND (screening OR "systematic review") | General search |
| B | (PABAK OR "Gwet's AC1") AND (LLM OR "automated screening") | Agreement metrics |
| C | ("two-stage" OR "hybrid pipeline") AND ("uncertain category") AND LLM | Hybrid architecture |
| D | ("stratified sampling") AND (LLM OR "AI screening") | Sampling design |

Raw yield: 893 records.

##### 2.B Federated multi-source search (MCP servers)

Fourteen sources queried in parallel: arXiv, PubMed, PMC, Europe PMC, CORE, OpenAlex, Semantic Scholar, Crossref, bioRxiv, medRxiv, DOAJ, BASE, Zenodo, HAL.

The same four themes (A-D), adapted to natural language for each endpoint. Raw yield: 226 records.

##### 2.C Citation chaining (Semantic Scholar API)

Forward and backward citation chasing from four seed studies identified in an initial exploratory pass (Elicit AI, 30 records):

| Seed | DOI |
|---|---|
| Ramchandani et al., 2025 | 10.7717/peerj-cs.2822 |
| Teperikidis et al., 2025/2026 (Synthesa AI) | 10.1101/2025.07.16.25331632 (preprint) / 10.1097/fjc.0[illegible] |
| Tao et al., 2025 (LitAutoScreener) | 10.34133/hds.0322 |
| Nawrath et al., 2026 | 10.3390/info17050501 |

Raw yield: 19 records. The chaining script collected incoming and outgoing citations of the four seeds but did not include the seeds themselves. The four seeds were added manually to the final set.

##### 2.D Manual addition

The four seeds in 2.C were added to the final pool with the tag manual_seed, because strategies 2.A and 2.B had not retrieved them.

#### 3. Deduplication

Two-step pipeline:

1. Exact DOI match (case-insensitive, normalised). 2. Fuzzy title match (Levenshtein distance, similarity at least 0.85) for records without a DOI, or with a duplicate DOI missed at step 1.

Result: 893 + 226 + 19 = 1,138 raw records, reduced to 508 unique records.

Two preprint/published duplicates below the automatic similarity threshold were found and corrected at quality control:

• Scherbakov et al. (arXiv 2409.04600 versus J Am Med Inform Assoc 2025;32:1071-1086): title similarity 0.82, below threshold.
• Janoudi et al. / Loon Lens (medRxiv 2024.09.06.24313186 versus Value Health 2025): title similarity 0.52 (titles substantially rewritten between preprint and published version); not detectable by title-only fuzzy match.

In both cases the peer-reviewed published version was kept and the duplicate preprint removed.

#### 4. Heuristic screening (DeepSeek V4 Pro)

The 508 records were screened automatically (DeepSeek API, temperature = 0, 8 parallel workers) as INCLUDE / EXCLUDE / UNCERTAIN for relevance to the gap analysis. This pass is not the clinical systematic-review screening.

Result: 84 INCLUDE, 401 EXCLUDE, 26 UNCERTAIN. The 110 INCLUDE + UNCERTAIN records went to structured extraction.

#### 5. Pre-extraction cleaning

Removed from the 110 records:

• 3 further preprint/published duplicates identified manually (Synthesa AI, and the two already listed in section 3, checked again in this batch).
• 6 records of general statistical literature on Cohen's kappa / PABAK / AC1 that are not screening-validation studies (for example the R package irrCAC, and general comparisons of agreement indices). These were moved to a separate reference file for the Methods section and were not counted as competing studies.

Final pool for structured extraction: 101 records.

#### 6. Structured extraction (8 dimensions)

Each of the 101 records was processed with the DeepSeek API (JSON mode, temperature = 0) to extract eight dimensions: reported agreement metrics, number of classification categories, pipeline architecture, validation sampling design, dataset prevalence or imbalance, application domain, direct overlap with the present design, and uncovered aspects (differential gap).

#### 7. Correction of truncated-abstract artefacts

Three records (MechaScreener, Auto-SLR, and the schema-constrained pipeline of Serretti) had received high scores in automatic extraction because the retrieval source (Crossref/MCP) returned only the first approximately 300 characters of the abstract. Manual check of the full text (PDFs retrieved directly) corrected the scores from 5/5, 5/5 and 3/5 to 1/5, 0/5 and 1/5. None of the three covers the five dimensions of the present design.

#### 8. Flow (simplified PRISMA style)

```
| Step | n |
|---|---|
| Structured search (findpapers) | 893 |
| Federated search (MCP, 14 sources) | 226 |
| Citation chaining (Semantic Scholar) | 19 |
| Raw total | 1,138 |
| After deduplication (exact DOI + fuzzy title >= 0.85) | 508 unique |
| Plus 4 seeds added manually | — |
| Heuristic screening, INCLUDE + UNCERTAIN | 110 |
| After cleaning (preprint/published duplicates + general statistical references) | 101 |
| Eight-dimension extraction (DeepSeek, JSON mode) | 101 |
| Manual full-text check of 6 candidates with score >= 2/5 | no study covers all 5 dimensions (maximum [illegible]
```

#### 9. Search date

Search run on 3 July 2026. Not updated afterwards. Studies published after that date are not included in the gap analysis.
