## Supplementary Materials (Files S1-S5) for "Validation of a two-stage automated screening pipeline for medical systematic reviews: a stratified agreement study with three independent human reviewers, using PABAK and Gwet’s AC1": File_S2_agreement_metrics.py

```
#!/usr/bin/env python3
"""
File S2: agreement_metrics.py
Reproducible calculation of inter-rater concordance metrics for title-abstract screening validation.
Calculates:
  - Percent observed agreement (Po)
  - Prevalence- and Bias-Adjusted Kappa (PABAK, q=3)
  - Gwet's First Agreement Coefficient (AC1)
  - Sensitivity and Specificity (with Wilson 95% CIs)
  - Work Saved over Sampling (WSS) and Number Needed to Read (NNR)
  - Jaccard index on INCLUDE sets
"""

import sys
import math
import pandas as pd
import numpy as np

def wilson_score_interval(k, n, confidence=0.95):
    if n == 0:
        return (0.0, 0.0)
    z = 1.95996  # 95%
    p = k / n
    denominator = 1 + z**2 / n
    centre_adjusted_probability = p + z**2 / (2 * n)
    adjusted_standard_deviation = math.sqrt((p * (1 - p) + z**2 / (4 * n)) / n)
    lower_bound = (centre_adjusted_probability - z * adjusted_standard_deviation) / denominator
    upper_bound = (centre_adjusted_probability + z * adjusted_standard_deviation) / denominator
    return (max(0.0, lower_bound), min(1.0, upper_bound))

def compute_pabak(po, q=3):
    """
    PABAK formula for q categories:
    PABAK = (q * Po - 1) / (q - 1)
    """
    return (q * po - 1.0) / (q - 1.0)

def compute_gwet_ac1(df, col1, col2, categories=('INCLUDE', 'EXCLUDE', 'UNCERTAIN')):
    """
    Compute Gwet's AC1 for 2 raters on categorical classifications.
    Pe = sum_{c} pi_c * (1 - pi_c) / (q - 1)
    where pi_c is the average proportion of category c assigned across both raters.
    AC1 = (Po - Pe) / (1 - Pe)
    """
    sub = df[[col1, col2]].dropna()
    n = len(sub)
    if n == 0:
        return 0.0, 0.0, 0.0
    po = (sub[col1] == sub[col2]).mean()
    q = len(categories)
    
    # marginal probabilities
    pi_sum = 0.0
    for c in categories:
        p1 = (sub[col1] == c).mean()
        p2 = (sub[col2] == c).mean()
        pi_c = (p1 + p2) / 2.0
        pi_sum += pi_c * (1.0 - pi_c)
        
    pe = pi_sum / (q - 1.0)
    if pe >= 1.0:
        ac1 = 1.0
    else:
        ac1 = (po - pe) / (1.0 - pe)
    return po, pe, ac1

def compute_pairwise_table(df, rater_cols, labels=('INCLUDE', 'EXCLUDE', 'UNCERTAIN')):
    results = []
    names = list(rater_cols.keys())
    for i in range(len(names)):
        for j in range(i + 1, len(names)):
            r1, r2 = names[i], names[j]
            c1, c2 = rater_cols[r1], rater_cols[r2]
            valid = df[[c1, c2]].dropna()
            n = len(valid)
            po, pe, ac1 = compute_gwet_ac1(valid, c1, c2, categories=labels)
            pabak = compute_pabak(po, q=len(labels))
            agreed = int(round(po * n))
            results.append({
                "Rater 1": r1,
                "Rater 2": r2,
                "n": n,
                "Agreed": agreed,
                "Po": f"{po*100:.1f}%",
                "PABAK": f"{pabak:.3f}",
                "AC1": f"{ac1:.3f}"
            })
    return pd.DataFrame(results)

def main():
    csv_file = "File_S4b_with_Berti_majority.csv"
    if len(sys.argv) > 1:
        csv_file = sys.argv[1]
        
    print(f"Loading validation data from: {csv_file}")
    df = pd.read_csv(csv_file)
    
    # Column mapping
    # Expected columns:
    # machine_decision (AI), human_decision_enrico (E.P.), human_decision_costazza (D.C.), 
    # human_decision_berti (Berti), majority_decision (Majority)
    col_map = {}
    for col in df.columns:
        cl = col.lower()
        if "machine" in cl or cl == "ai":
            col_map["AI"] = col
        elif "enrico" in cl or "ep" in cl:
            col_map["E.P."] = col
        elif "costazza" in cl or "dc" in cl:
            col_map["D.C."] = col
        elif "berti" in cl or "ppb" in cl:
            col_map["Berti"] = col
        elif "majority" in cl:
            col_map["Majority"] = col
            
    print(f"Detected columns: {col_map}")
    
    # 1. Overall Pairwise Table
    print("\n--- TABLE 3: Overall Pairwise Concordance (q=3) ---")
    pw = compute_pairwise_table(df, col_map)
    print(pw.to_string(index=False))
    
    # 2. Majority distribution
    if "Majority" in col_map:
        maj_col = col_map["Majority"]
        print("\n--- Majority Distribution (n=185) ---")
        print(df[maj_col].value_counts(dropna=False))
        
    # 3. Include sets and Jaccard
    print("\n--- INCLUDE Sets and Jaccard Overlap ---")
    include_sets = {}
    id_col = "record_id" if "record_id" in df.columns else df.columns[0]
    for rater, c in col_map.items():
        include_ids = set(df[df[c] == "INCLUDE"][id_col])
        include_sets[rater] = include_ids
        print(f"{rater} INCLUDE count: {len(include_ids)} -> IDs: {sorted(list(include_ids))[:10]}...")
        
    print("\nPairwise Jaccard on INCLUDE sets:")
    names = list(include_sets.keys())
    for i in range(len(names)):
        for j in range(i + 1, len(names)):
            s1, s2 = include_sets[names[i]], include_sets[names[j]]
            inter = len(s1 & s2)
            union = len(s1 | s2)
            jacc = inter / union if union > 0 else 0.0
            print(f"{names[i]} vs {names[j]}: {inter}/{union} = {jacc:.3f}")

if __name__ == "__main__":
    main()
```
